# Serial interval of novel coronavirus (COVID-19) infections

**DOI:** 10.1101/2020.02.03.20019497

**Authors:** Hiroshi Nishiura, Natalie M. Linton, Andrei R. Akhmetzhanov

## Abstract

**Objective:** To estimate the serial interval of novel coronavirus (COVID-19) from information on 28 infector-infectee pairs.

**Methods:** We collected dates of illness onset for primary cases (infectors) and secondary cases (infectees) from published research articles and case investigation reports. We subjectively ranked the credibility of the data and performed analyses on both the full dataset (*n*=28) and a subset of pairs with highest certainty in reporting (*n*=18). In addition, we adjusting for right truncation of the data as the epidemic is still in its growth phase.

**Results:** Accounting for right truncation and analyzing all pairs, we estimated the median serial interval at 4.0 days (95% credible interval [CrI]: 3.1, 4.9). Limiting our data to only the most certain pairs, the median serial interval was estimated at 4.6 days (95% CrI: 3.5, 5.9).

**Conclusions:** The serial interval of COVID-19 is shorter than its median incubation period. This suggests that a substantial proportion of secondary transmission may occur prior to illness onset. The COVID-19 serial interval is also shorter than the serial interval of severe acute respiratory syndrome (SARS), indicating that calculations made using the SARS serial interval may introduce bias.

**Highlights:**

- The serial interval of novel coronavirus (COVID-19) infections was estimated from a total of 28 infector-infectee pairs.
- The median serial interval is shorter than the median incubation period, suggesting a substantial proportion of pre-symptomatic transmission.
- A short serial interval makes it difficult to trace contacts due to the rapid turnover of case generations.

## Introduction

The epidemic of novel coronavirus (COVID-19) infections that began in China in late 2019 has rapidly grown and cases have been reported worldwide. An empirical estimate of the serial interval—the time from illness onset in a primary case (infector) to illness onset in a secondary case (infectee)—is needed to understand the turnover of case generations and transmissibility of the disease [1]. Estimates of the serial interval can only be obtained by linking dates of onset for infector-infectee pairs, and these links are not easily established. A recently published epidemiological study used contact tracing data from cases reported in Hubei Province early in the epidemic to estimate the mean serial interval at 7.5 days [2], which is consistent with the 8.4-day mean serial interval reported for severe acute respiratory syndrome (SARS) from Singaporean household contact data [3]. However, there were only six infector-infectee pairs in this dataset, and sampling bias may have been introduced to the variance and mean. To further assess the serial interval of COVID-19 infections we compiled a dataset of 28 publicly shared infector-infectee pairs and calculated the serial interval from these data.

## Materials and Methods

We scanned publicly available information published in research articles and quoted from official reports of outbreak investigations to obtain our dataset. The date of illness onset was defined as the date on which a symptom relevant to COVID-19 infection appeared and was determined by the reporting governmental body. We subjectively ranked the credibility of the ascertained pairs into “certain” and “probable,” where the former was used for pairs and dates of illness onset were clearly defined in an academic article and the latter was applied to pairs and dates of illness onset that were clearly defined but quoted from outbreak investigation reports. Estimates were obtained for certain and probable pairs combined (*n*=28) as well as for the certain pairs alone (*n*=18).

The interval censored data were handled in units of days. We employed a Bayesian approach with doubly interval censored likelihood to obtain estimates of the serial interval [4]:

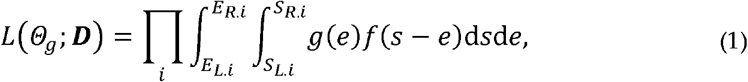

where *i* represents the identity of each pair, *E*(*R,L*) is the interval for symptom onset of the infector and *S*(*R,L*) is the interval for symptom onset of the infectee. Here, *g*(.) is the probability density function (p.d.f.) of exposure following a uniform distribution and *f*(.) is the p.d.f. of the serial interval, assumed to be governed by three different distributions—lognormal, gamma, and Weibull. We sampled the posterior distributions using CmdStan version 2.22.1 (http://github.com/aakhmetz/nCoVSerialInteval2020).

As the epidemic will continue to grow beyond our data collection cutoff point of 12 February 2020, it is possible that the naïve likelihood (1) underestimates the serial interval as sampling during the early stage of the epidemic preferentially excludes infector-infectee pairs with longer serial intervals. We adjusted for this selection bias—called right truncation—in our model. The alternative p.d.f. that accounts for right truncation during the exponential growth phase of the epidemic is written as:

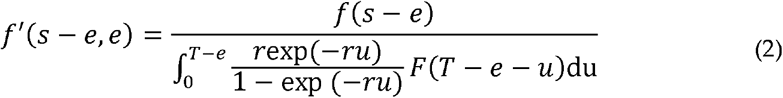

where *r* is the exponential growth rate estimated at 0.14 [5] and *T* is the latest time of observation (12 February 2020). The widely applicable information criterion (WAIC) was used to compare between distributions and the model with the minimal WAIC value was selected as the best-fit model for each set of estimates with and without right truncation.

## Results

We were able to obtain data on 28 infector-infectee pairs (see Supplementary Table). Of these, 12 pairs were family clusters. Accounting for right truncation and analyzing all pairs, the model using the lognormal distribution was selected as the best-fit model (WAIC=224.0) The median serial interval was estimated at 4.0 days (95% credible interval [CrI]: 3.1, 4.9) while the mean and standard deviation (SD) of the serial interval were estimated at 4.7 days (95% CrI: 3.7, 6.0) and 2.9 days (95% CrI: 1.9, 4.9), respectively. Without truncation, the model using the lognormal distribution was also the best-fit model (WAIC=128.0) with the median serial interval was estimated at 3.9 days (95% CrI: 3.1, 4.8).

Limiting our dataset to only certain observations, the median serial interval of the best-fit Weibull distribution model was estimated at 4.6 days (95% CI: 3.5, 5.9) with a mean and SD of 4.8 days (95% CrI: 3.8, 6.1) and 2.3 days (95% CrI: 1.6, 3.5), respectively. Without truncation, the best-fit model used the lognormal distribution and estimated the median serial interval at 4.1 days (95% CrI: 3.2, 5.0). Figure 1 shows the best-fit distributions overlaid with a published distribution of the SARS serial interval [4].

**Figure 1.**
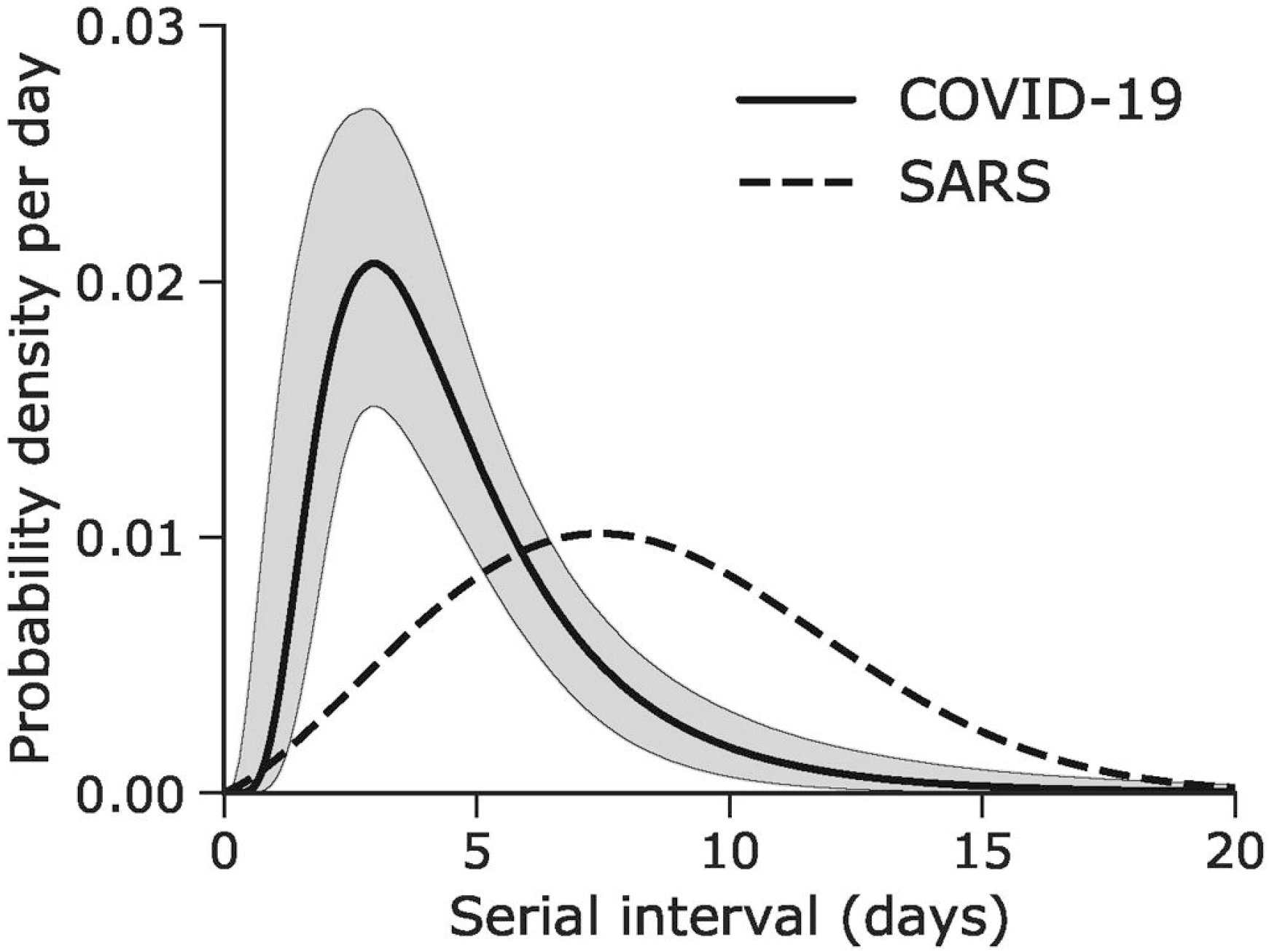
Serial interval of novel coronavirus (COVID-19) infections. The solid line shows the estimated serial interval distribution of COVID-19 infections using the best-fit lognormal distribution with right truncation. A distribution based on a published estimate of the serial interval for severe acute respiratory syndrome [3] is overlaid as a dashed line for comparison.

## Discussion

Our estimate of the median serial interval as 4.0 days indicates that COVID-19 infection leads to rapid cycles of transmission from one generation of cases to the next. The shorter serial interval compared to SARS implies that contact tracing methods must compete against the rapid replacement of case generations, and the number of contacts may soon exceed what available healthcare and public health workers are able to handle. The difference between these distributions suggests that using serial intervals estimates from SARS data will result in overestimation of the COVID-19 basic reproduction number.

More importantly, the estimated median serial interval is shorter than the preliminary estimates of the mean incubation period (approximately 5 days) [3,6]. As illustrated in Figure 2, when the serial interval is shorter than the incubation period, pre-symptomatic transmission is likely to have taken place and may even occur more frequently than symptomatic transmission. A substantial proportion of secondary transmission occurring before illness onset indicates that many transmissions cannot be prevented solely through isolation of symptomatic cases, as by the time contacts are traced they may have already become infectious themselves and generated secondary cases [7].

**Figure 2.**
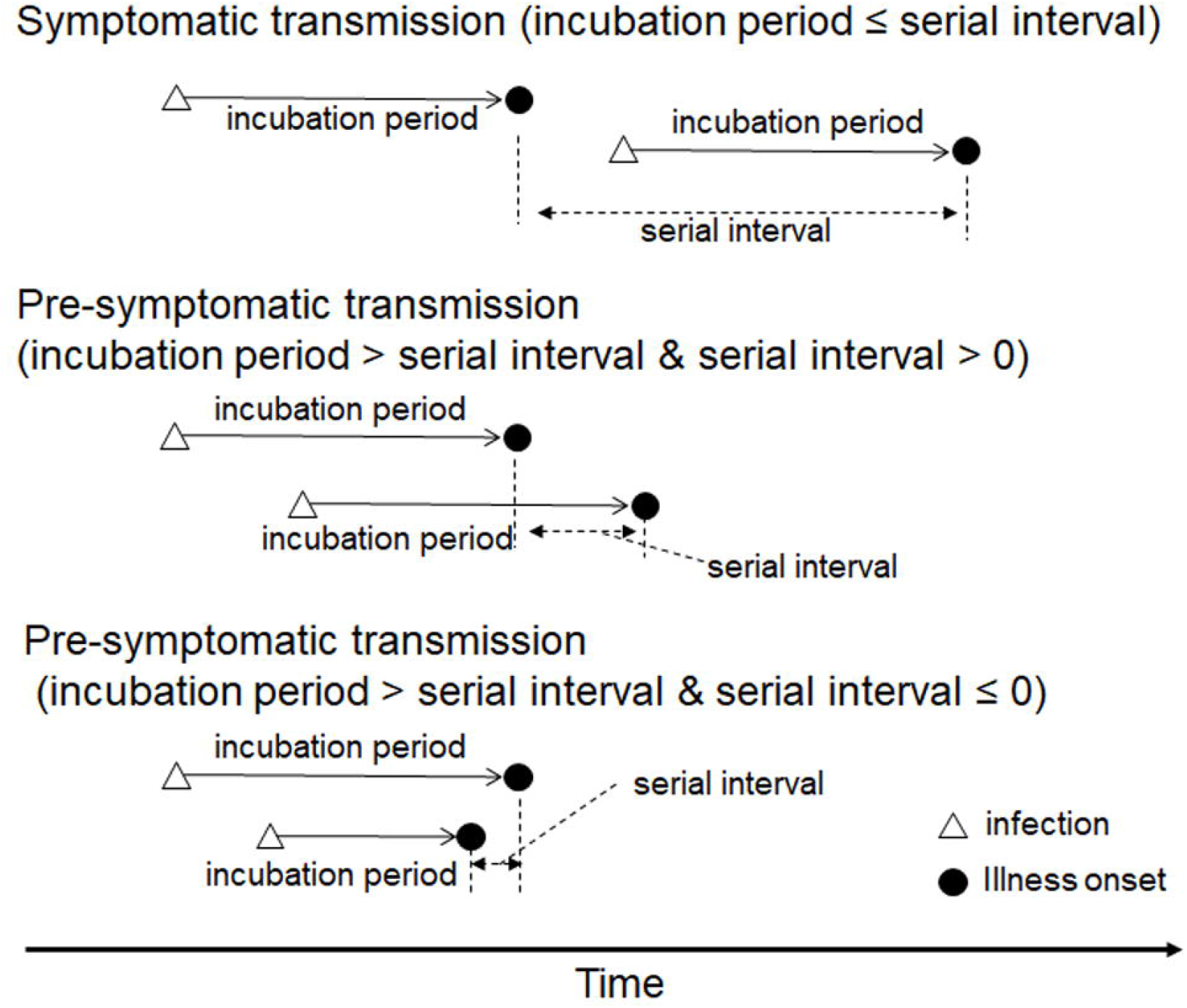
The relationship between the incubation period and serial interval. If the transmission takes place during the symptomatic period of the primary case, the serial interval is longer than the incubation period. However, this relationship can be reversed when pre-symptomatic transmission takes place (the secondary case may even experience illness onset prior to onset in their infector).

Correct ascertainment of dates of illness onset is critical to the calculation of the serial interval. Considering the overall mild nature of the infection [8] it is possible that different reporting jurisdictions have different criteria for determining what qualifies as illness onset for COVID-2019 cases, which is a potential bias we are unable to account for. However, the present study addresses the issue of data quality of the reported pairs in two ways. First, our data include the updated information from a recent report of pre-symptomatic transmission in Germany [9] where it was later found that the primary case was already symptomatic while in contact with persons who later became infected (Supplementary Material in [9]). Second, classification of the credibility of the data and comparing analyses including and excluding less certain (but nonetheless highly probable) pairs allowed us to determine that our results using all pairs (and therefore a greater sample size) did not differ significantly from the results using only the most credible data.

In conclusion, we have estimated the median serial interval of COVID-19 at 4.0 days, which is shorter than the disease’s median incubation period indicating that rapid cycles of transmission and substantial pre-symptomatic transmissions are occurring. Thus, containment via case isolation alone is likely to be very challenging.

## Data Availability

The data can be obtained from Supplementary Table.

## Acknowledgments

H.N. received funding support from Japan Agency for Medical Research and Development [grant number: JP18fk0108050] the Japan Society for the Promotion of Science (JSPS) Grants-in-Aid for Scientific Research (KAKENHI in Japanese abbreviation) grant nos. 17H04701, 17H05808, 18H04895 and 19H01074, and the Japan Science and Technology Agency (JST) Core Research for Evolutional Science and Technology (CREST) program [grant number: JPMJCR1413]. NML received a graduate study scholarship from the Ministry of Education, Culture, Sports, Science and Technology, Japan. The funders had no role in study design, data collection and analysis, decision to publish, or preparation of the manuscript.

